# Major Cardiovascular Event Risk of Advanced Therapies in Inflammatory Bowel Diseases: Systematic Review and Meta-Analysis

**DOI:** 10.64898/2026.01.26.26344897

**Authors:** Amirah H. Alnahdi, Chelsea Salmon, Mikael Svensson, Naueen Chaudhry, Ellen M. Zimmermann, Tianze Jiao

**Affiliations:** Department of Pharmaceutical Outcomes and Policy, College of Pharmacy, University of Florida, Gainesville, Florida, USA; Center for Drug Evaluation and Safety (CoDES), University of Florida, Gainesville, Florida, USA; Therapeutic Affairs, Ministry of Health, Riyadh, Saudi Arabia; Division of Gastroenterology, Hepatology & Nutrition, Department of Medicine, University of Florida, Gainesville, Florida, USA; School of Public Health & Community Medicine, University of Gothenburg, Sweden

**Author notes:** **Corresponding author:** Tianze Jiao, PhD, Assistant Professor, Department of Pharmaceutical Outcomes & Policy College of Pharmacy, University of Florida. **Contact details:** Amirah H. Alnahdi, Chelsea Salmon, Mikael Svensson, Naueen Chaudhry, Ellen M. Zimmermann.

**Keywords:** Inflammatory Bowel Disease, Advanced Therapies, Major Adverse Cardiovascular Events

## Abstract

**Background:** Patients with chronic immune-mediated disorders (IMIDs), including inflammatory bowel disease (IBD), are at increased risk of cardiovascular disease. While advanced therapies show cardioprotective effects in other IMIDs, their impact on major adverse cardiovascular events (MACE) in IBD remains unclear. We conducted a meta-analysis of randomized controlled trials (RCTs) and observational studies evaluating MACE risk with advanced therapies in IBD.

**Methods:** Systematic search of PubMed, Embase and Cochrane Central identified 43 high-quality studies (36 RCTs,7 observational studies) published between 2002 and 2024. Primary analyses estimated odds ratios (OR) for MACE comparing advanced therapy to placebo, with secondary analyses stratified studies by drug class and length of follow-up.

**Results:** Placebo-controlled RCTs showed a nonsignificant trend toward reduced MACE risk (OR: 0.60; 95% CI: 0.24–1.51), with similar findings after continuity correction for zero-event studies (OR: 0.87; 95% CI: 0.45–1.68). Class-specific trends suggested lower MACE risk with IL-12/IL-23 inhibitors (OR: 0.35; 95% CI: 0.05–2.21), JAK inhibitors (OR: 0.57; 95% CI: 0.16–2.06), and a potential increase with anti-TNF agents (OR: 3.04; 95% CI: 0.31–29.47), though none reached statistical significance. Long-term follow-up studies showed consistent findings. Observational studies suggested lower MACE risk for anti-TNF therapies (OR: 0.29; 95% CI: 0.21–0.40), but not for IL-12/IL-23 (OR: 4.41; 95% CI: 0.49–39.28) or JAK inhibitors (OR: 1.57; 95% CI: 0.86–2.84).

**Conclusion:** Advanced therapies did not demonstrate a clear increase or decrease in cardiovascular risk in IBD. The discrepancies between RCTs and observational studies underscore the urgent need for rigorous-designed observational research with long-term follow-up to evaluate the real-world impact of advanced therapies on MACE risk.

**Summary:** This study evaluated cardiovascular safety of advanced therapies in inflammatory bowel disease. Findings showed no clear signal of decreased major cardiovascular risk compared with conventional treatment, highlighting the need for continued monitoring through long-term and real-world evidence.

**Key Messages:**

- What is already known? Patients with IBD are at increased risk of cardiovascular events, and the impact of advanced therapies on this risk remains uncertain.
- What is new here? This meta-analysis integrates data from randomized and observational studies and found no significant association between advanced therapies and MACE.
- How can this study help patient care? Findings show no clear signal of decreased cardiovascular risk with advanced IBD therapies, though continued evaluation is warranted.

## Background

Inflammatory bowel diseases (IBD), encompassing Crohn’s disease (CD) and ulcerative colitis (UC), are chronic, immune-mediated inflammatory disorders (IMIDs) impacting over 0.3% of the global population^1^. The prevalence of IBD has risen markedly over recent decades, with cases increasing from 3.3 million in 1990 to 4.9 million worldwide in 2019, reflecting the growing global burden of this disease^2^. In the United States, IBD is a significant public health concern, with recent epidemiological studies underscoring its substantial prevalence, approximately 0.7% of Americans are diagnosed with IBD ^3^.

IBD treatment has evolved significantly in the past two decades, and an expanding number of therapeutic options available to manage symptoms and induce clinical and endoscopic remission. With advancements in therapies that target inflammatory processes, treatment objectives include symptom control and mucosal healing, ultimately improving long-term health outcomes for patients with IBD.^4,5^. Advanced therapies for IBD include antibodies to, or inhibitors of, relevant inflammatory targets. These include anti-tumor necrosis factor therapies (anti-TNF), anti-interleukin (IL) treatments (anti-IL-23, anti-IL-12/23), integrin receptor antagonists, janus kinase (JAK) inhibitors, and sphingosine-1-phosphate receptor (S1PR) modulators. Not surprisingly, these therapies are effective in several IMIDs including rheumatoid arthritis, poriasis, psoriatic arthritis, and ankylosing spondylitis.

Many of the same inflammatory pathways are important to our modern understanding of atherogenesis. Patients with IMIDs have an increased risk of cardiovascular disease and an elevated risk of major adverse cardiovascular events (MACE). Notably, the elevated risk of ischemic heart disease in individuals with IBD has been substantiated by two meta-analyses published in 2014, demonstrating an odds ratio (OR) of 1.19 (95% CI: 1.08–1.31)^6^, and a relative risk (RR) of 1.35 (95% CI: 1.19-1.52)^7^. The increased risk of cardiovascular disease is also seen in patients with other IMIDs, such as rheumatoid arthritis (RA) and axial spondyloarthritis (axSpA): increased risk of cardiovascular death (hazard ratio [HR]1.41, 95% CI: (1.35-1.49) and 1.40, 95% CI: (1.21-1.62), respectively)^8^.

Some advanced therapies used in IBD and other IMIDs have demonstrated cardioprotective effects. Previous meta-analyses have indicated that anti-TNF therapies are associated with reduced cardiovascular risk in RA patients with a relative risk of 0.70, 95% CI: (0.54 -0.90)^9^. Similarly, a retrospective cohort study using a health plan database found that in psoriasis patients without major cardiovascular disease [MACE], anti-TNF therapy was associated with a significantly lower risk of MACE (HR: 0.80, 95% CI: (0.66–0.98)) compared to topical therapy after adjusting for cardiovascular risk factors by propensity score^10^. Indeed, inflammatory markers commonly elevated in IBD, including C-reactive protein (CRP), interleukin-6 (IL-6), IL-1, IL-8, and tumor necrosis factor-alpha (TNF-α), have demonstrated predictive value for future cardiovascular (CV) events^11^. Moreover, a reduction in serum levels of IL-6 and TNF-α has been correlated with improvements in cardiovascular outcomes^12^.

Although the cardioprotective effects of these therapies have been demonstrated in other IMIDs and are pathologically plausible for IBD, the effect of therapy on cardiovascular risk for patients with IBD remains an open question. One recently published meta-analyses, Shehab et al.(2023)^13^, addressed cardiovascular risk through randomized controlled trials (RCTs). They found that during the induction period the use of advanced therapy was associated with approximately 30% risk reduction in MACE, for both CD and UC patients [CD, OR: 0.69, 95% CI: (0.26-1.82); UC, OR: 0.61, 95% CI: (0.29-1.28)]. During the maintenance phase, the risk reduction varied, ranging from 10% for CD to 40% for UC [CD, OR: 0.91, 95% CI: (0.28-2.90); UC, OR: 0.59, 95% CI: (0.22-1.60)]. In our current study, we enhanced our systematic review and meta-analysis to include both RCTs and observational studies, focusing on patients receiving advanced treatments. For the first time, we evaluate the effect of advanced therapy for IBD by drug class for the anti-TNFs, JAK inhibitors, anti-interleukin (IL) treatments (anti-IL-23, anti-IL-12/23), and integrin receptor antagonists. We used statistical methods to address important biases induced by zero or low events in some trials and used methods to address mismeasured cardiovascular events. Most significantly, we compared the differences in risk assessment based on RCT vs. observational studies, highlighting the biases inherent in either strategy.

## Methods

### Data Sources and Search Strategy

A systematic literature search was conducted to identify relevant RCTs and observational studies, encompassing both cohort and case-control designs, and was registered in PROSPERO (Registration ID: [CRD420251162343]) while adhering to the PRISMA 2020 reporting guidelines. The main research databases were Embase and PubMed from inception to February 15, 2024. Key search terms were derived from a review of published literature in the field, with the complete list provided in the Supplementary materials.

The screening protocol was implemented in a two-stage approach utilizing the Covidence systematic review software^14^: initial screening of titles and abstracts, followed by a comprehensive evaluation of full-text articles. Two authors (A.A. and S.C.) independently performed this process, discrepancies arising during the data extraction process were resolved through a third author (F.D), and consultation with a senior author (T.J.) when necessary. To ensure comprehensive coverage, and identify additional eligible studies, reference lists of included articles and relevant systematic reviews were manually examined. The search strategy and study selection process are illustrated in (Figure 1).

**Figure 1:**
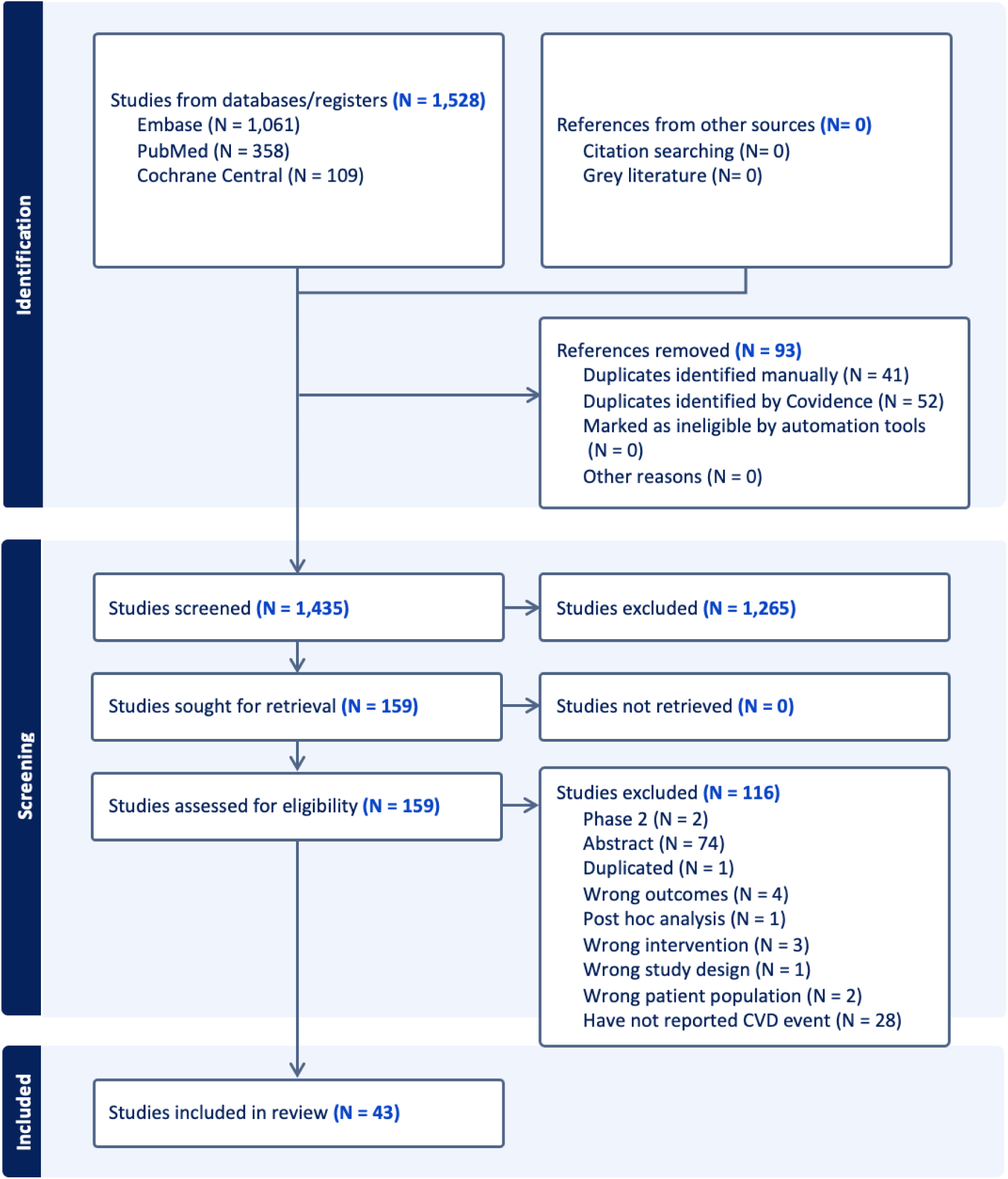
Flow diagram of the search strategy and study selection process.

### Selection Criteria

Studies were included if the following predetermined criteria were met: study design included RCTs, cohort studies, or case-control studies; assessed advance therapies for treating IBD and measured or mentioned MACE. For RCTs that have multiple time points for randomization, such as at the beginning of induction and maintenance period, we extracted data respectively from related time point of randomization till the occurrence of outcome, end of study, or the next randomization point, which ever occurred first. For RCTs reporting outcomes at multiple time points, we extracted data with the longest follow-up period that maintained the treatment assigned from randomization in the primary analysis. For RCTs that lost their randomization feature after a certain point—when all patients began receiving the same drug according to the protocol, we extracted data right before that time point for the RCT analyses. Events occurred thereafter were included in the long-term follow-up analyses. No restrictions were applied in terms of follow-up duration. Studies that did not examine patients with IBD or did not mention MACE events were excluded. These selection criteria were designed to ensure the inclusion of studies relevant to safety issues associated with advanced therapies for IBD, with a specific focus on cardiovascular events.

### Data Extraction and Quality Assessment

Data extraction was independently performed by two authors (A.A. and S.C.) using standardized forms. The extracted information encompassed: the first author’s surname, year of publication, study design, sample size, underlying condition (Crohn’s disease, ulcerative colitis, or indeterminate colitis), study duration, population characteristics, smoking status, comorbidities, exposure (drug and dose), concomitant advanced therapies, and reported outcomes. A detailed overview of the extracted studies is summarized in Supplementary Table S1. The primary efficacy outcome was MACE, a composite outcome of the myocardial infarction, unstable angina, cerebrovascular accident, cardiovascular death, or heart failure^15^.Any discrepancies during the data extraction process were resolved through consultation with the senior author (T.J.). The quality of included RCTs was assessed using the Cochrane Risk-of-Bias Tool for Randomized Trials (RoB2), while observational studies were evaluated using the Risk-of-Bias in Non-randomized Studies of Interventions (ROBINS-I) tool. This approach ensured a comprehensive and standardized assessment of potential biases across different studies ^16,17^. Detailed methodological information regarding the risk of bias assessment is provided in the Supplementary Tables S2 and S3.

### Statistical Analysis

The cardiovascular events in all studies were captured and reported in the odds ratio (OR). Data synthesis was conducted using the Mantel-Haenszel method, applying a random-effects model to provide more conservative estimates when inter-study variability was present. Results are presented as pooled odds ratios (OR) with 95% confidence intervals (CI). Inter-study heterogeneity was quantified using the I² statistic.

Our primary analysis focused on direct comparisons between advanced therapies and placebo in RCTs. To identify potential biases arising from studies with no events, we employed the continuity correction method using the reciprocal of the opposite treatment arm size^18^, which addresses the imbalance sample size between the exposure and control group. For our secondary analysis, subgroup analyses were conducted, stratified by pharmacological medication class and dosage regimens, to explore potential differences on the risk of MACE. Meta-regression analyses were applied to studies comparing advanced therapies with active comparators, including other advanced therapies, immunomodulators, and corticosteroids. Finally, data from long-term follow-up studies of RCTs and observational studies were incorporated, respectively, allowing for the assessment of result robustness across diverse study designs and follow-up durations. Publication bias was evaluated using a combination of visual inspection of funnel plots and quantitative assessment with Egger’s regression asymmetry test (detail in Supplementary Figure S.4).

All statistical analyses were performed using R software (version 2023.06.2+561), with a P-value <0.05 considered statistically significant.

## Result

### Study Characteristics

A total of 1,419 citations were identified through the systematic search strategy, with 159 articles undergoing thorough full-text examination. Ultimately, 43 eligible studies were included: 27 RCTs, 9 long-term follow-up (LTF), and 7 observational studies^(19–61)^. These studies encompassed 17 and 22 focusing on patients with CD and UC, respectively, and 4 involving both CD and UC patients. The publication dates ranged from 2007 to 2024. Baseline characteristics of the included studies are presented in Supplementary Table S1.

### Primary Analysis

The primary analysis included 19 placebo-controlled studies evaluating various advance therapies^(19–37)^: Anti-TNF agents (adalimumab [n=6], certolizumab pegol [n=1], and infliximab [n=1]), a Sphingosine-1-Phosphate Receptor (S1PR) modulator (ozanimod [n=1]), an Interleukin-23 (IL-23) inhibitor (risankizumab [n=2]), Janus kinase (JAK) inhibitors (tofacitinib [n=1], upadacitinib [n=2], and filgotinib [n=1]), an Interleukin-12/23 (IL-12/23) inhibitor (ustekinumab [n=2]), and an Integrin receptor antagonist (vedolizumab [n=1], and etrolizumab[n=1]). The pooled analysis of MACE showed an OR of 0.60 (95% CI: 0.24–1.51, p=0.78) (Figure 2). After applying a continuity correction, there was still no statistically significant difference in MACE risk between patients receiving advanced therapies and placebo (OR 0.87, 95% CI: 0.45–1.68) (Figure 3).

**Figure 2:**
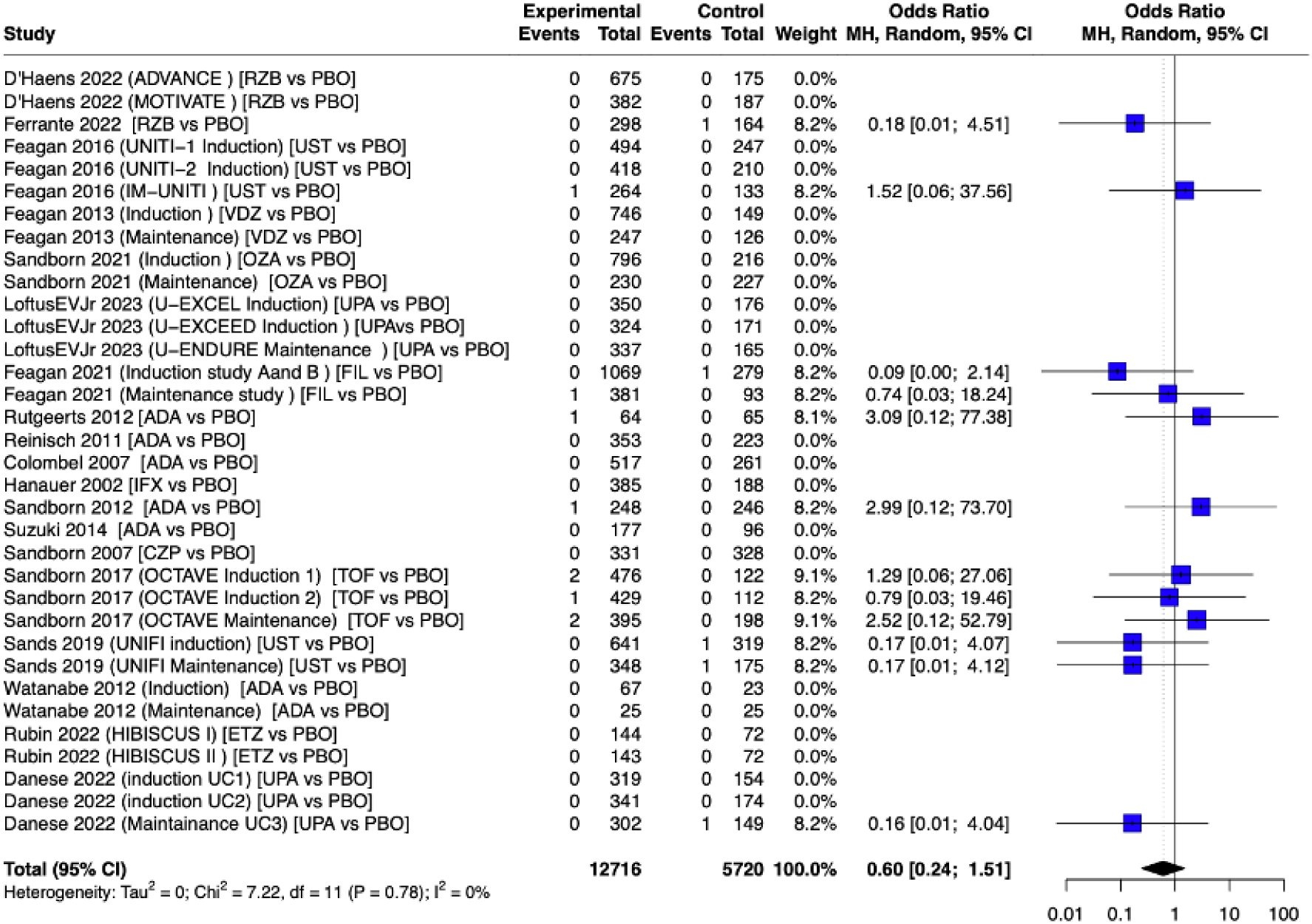
Forest Plot of Pooled Analysis for the MACE (advance therapies vs. placebo).

**Figure 3:**
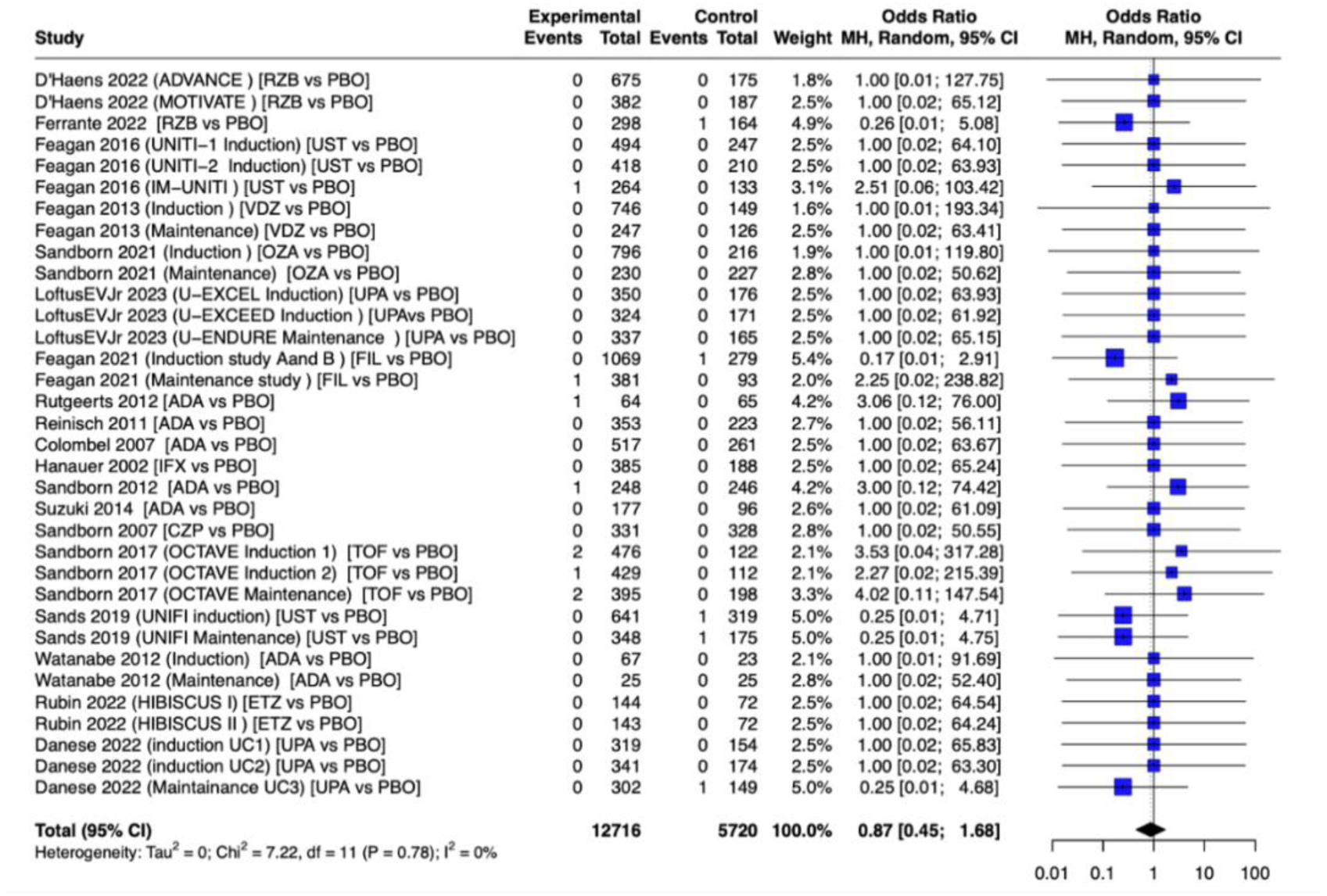
Forest Plot of Pooled Analysis for the MACE with continuity correction (advance therapies vs placebo).

### Secondary Analysis I: Stratified by Pharmacological Class

Stratification by pharmacological class revealed class-specific trends (Figure 4). IL-12/23 inhibitors (OR 0.35, 95% CI: 0.05–2.21) and JAK inhibitors (OR 0.57, 95% CI: 0.16–2.06) showed point estimates below 1, while anti-TNF therapies (OR 3.04, 95% CI: 0.31–29.47) showed a potential increased MACE risk. However, none of these associations were statistically significant.

**Figure 4.**
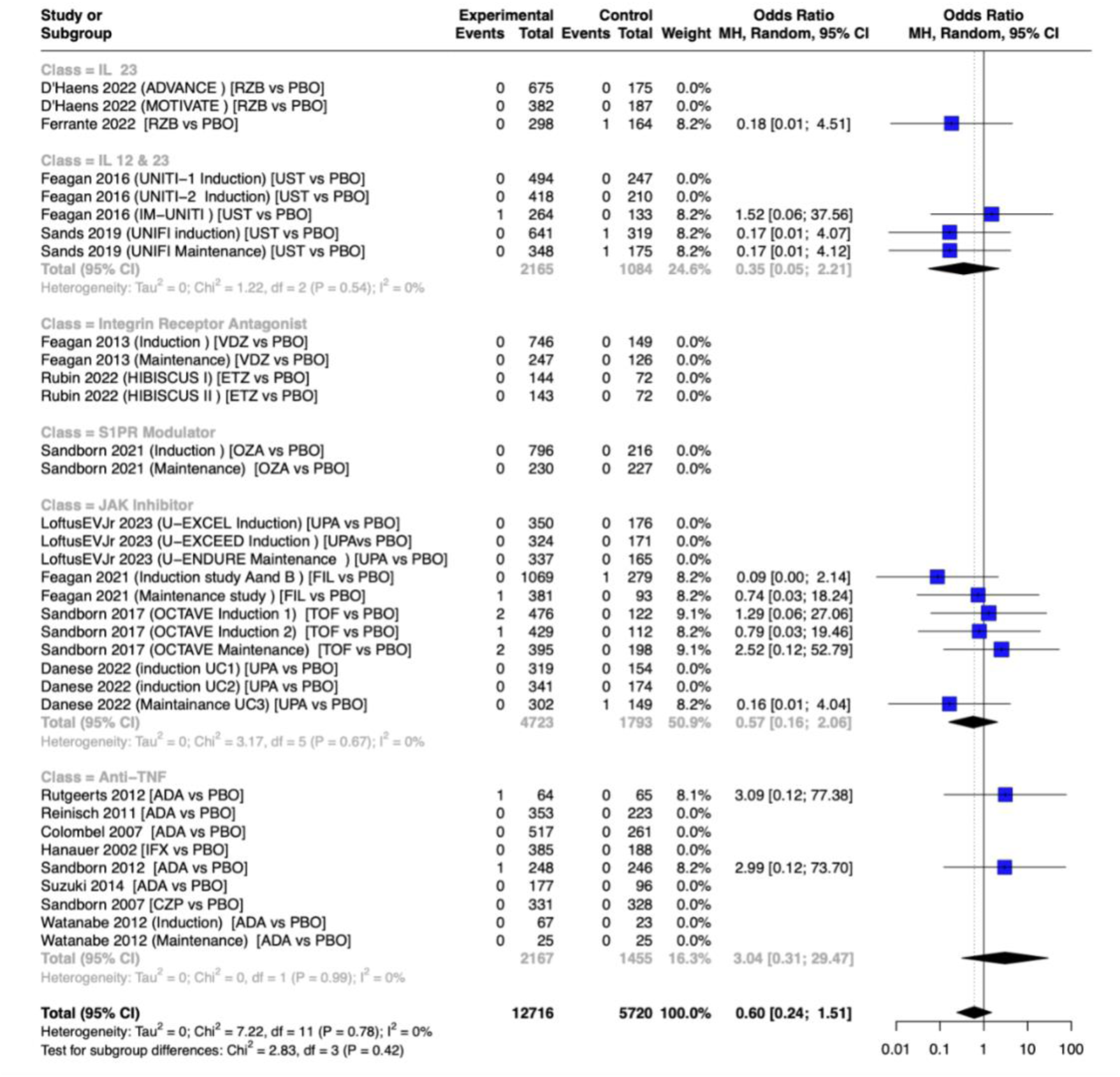
Forest Plot of Pooled Analysis for the MACE stratified by Pharmacological Class.

### Secondary Analysis II: Comparisons Between Higher and Lower Doses

Secondary analyses comparing higher and lower doses ^19,20,34,35,37–41,22,24,25,27–29,31,33^ yield a potential dose-dependent effects, with an overall (OR 0.61, 95% CI (0.16-2.36)), suggesting a possible trend toward reduced MACE risk with higher doses (Figure 5). However, these results did not reach statistical significance due to the small sample size and low event rate. After applying the continuity correction, the overall OR moved closer to 1 (0.90, 95% CI: (0.42-1.96)). Additional subgroup analyses comparing individual advance therapies with active comparators ^36,42–45^, moderator, and sensitivity analyses are provided in the Supplementary Materials, none of the associations was statistically significant.

**Figure 5.**
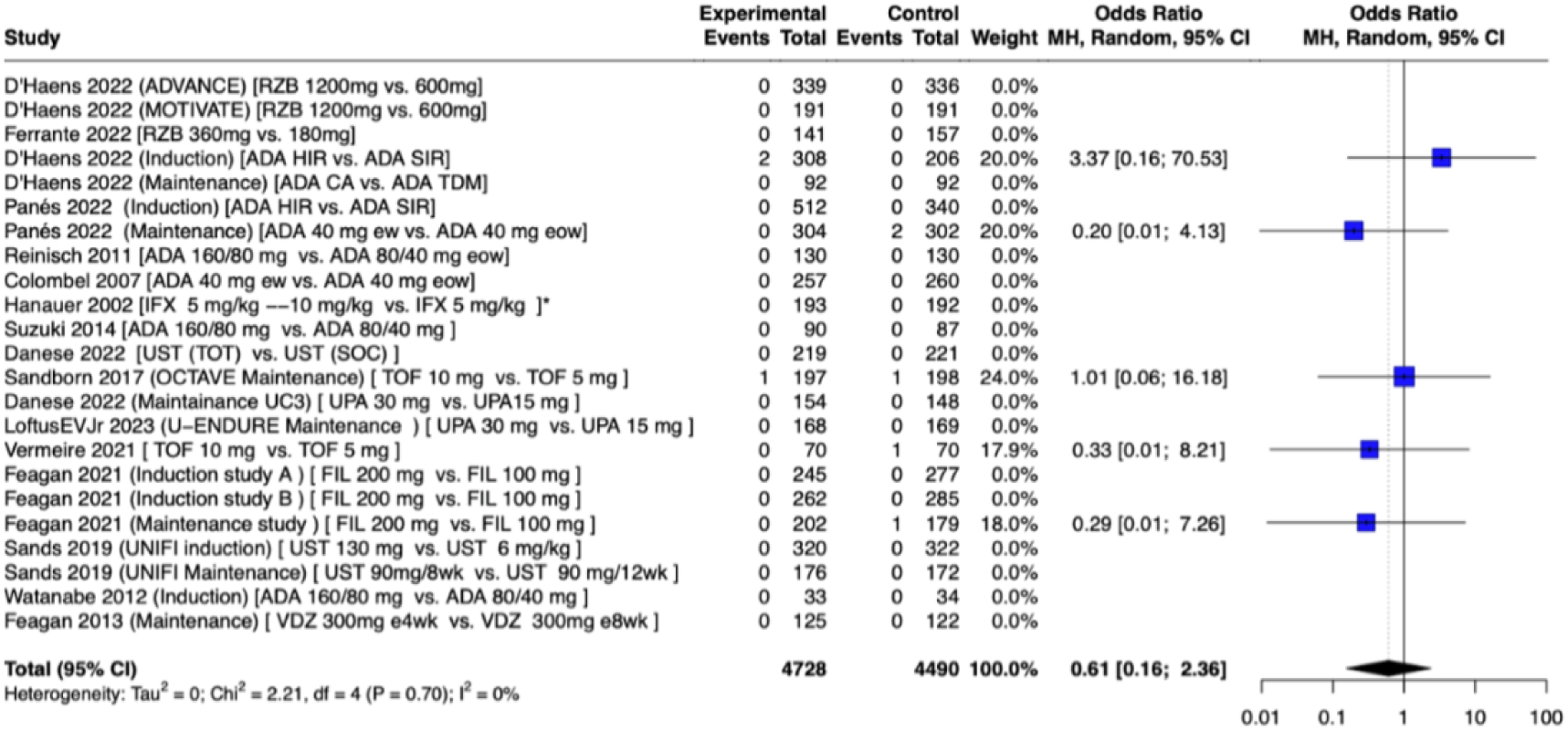
Forest Plot of Pooled Analysis that Compared the Dose of Same Medication for the MACE (high dose vs. low dose).

### Secondary Analysis III: Long-term Follow-up (LTF) and Observational Studies

LTF Studies ^46–54^ and observational studies^55–61^ showed consistent results with our primary analysis: advanced therapies have been associated with statistically insignificant reductions in MACE risk (OR 0.51, 95% CI: 0.19–1.35 for LTF studies; OR 0.81, 95% CI: 0.34–1.93 for observational studies) (Figures 6,7). However, these effects were not statistically significant, and the wide confidence intervals indicate uncertainty.

**Figure 6.**
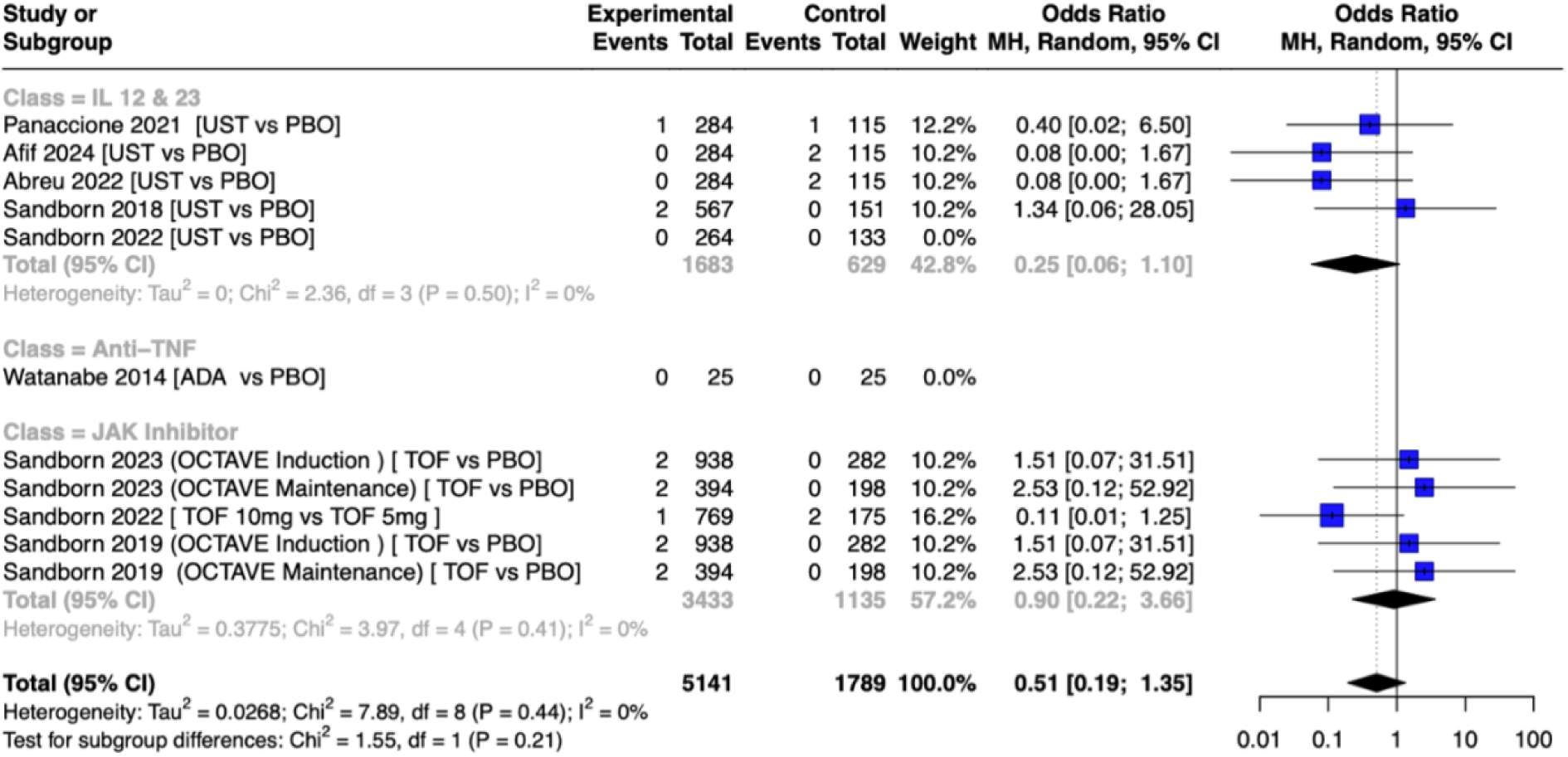
Forest Plot of Pooled Analysis for the MACE with Long-term Follow-up.

**Figure 7.**
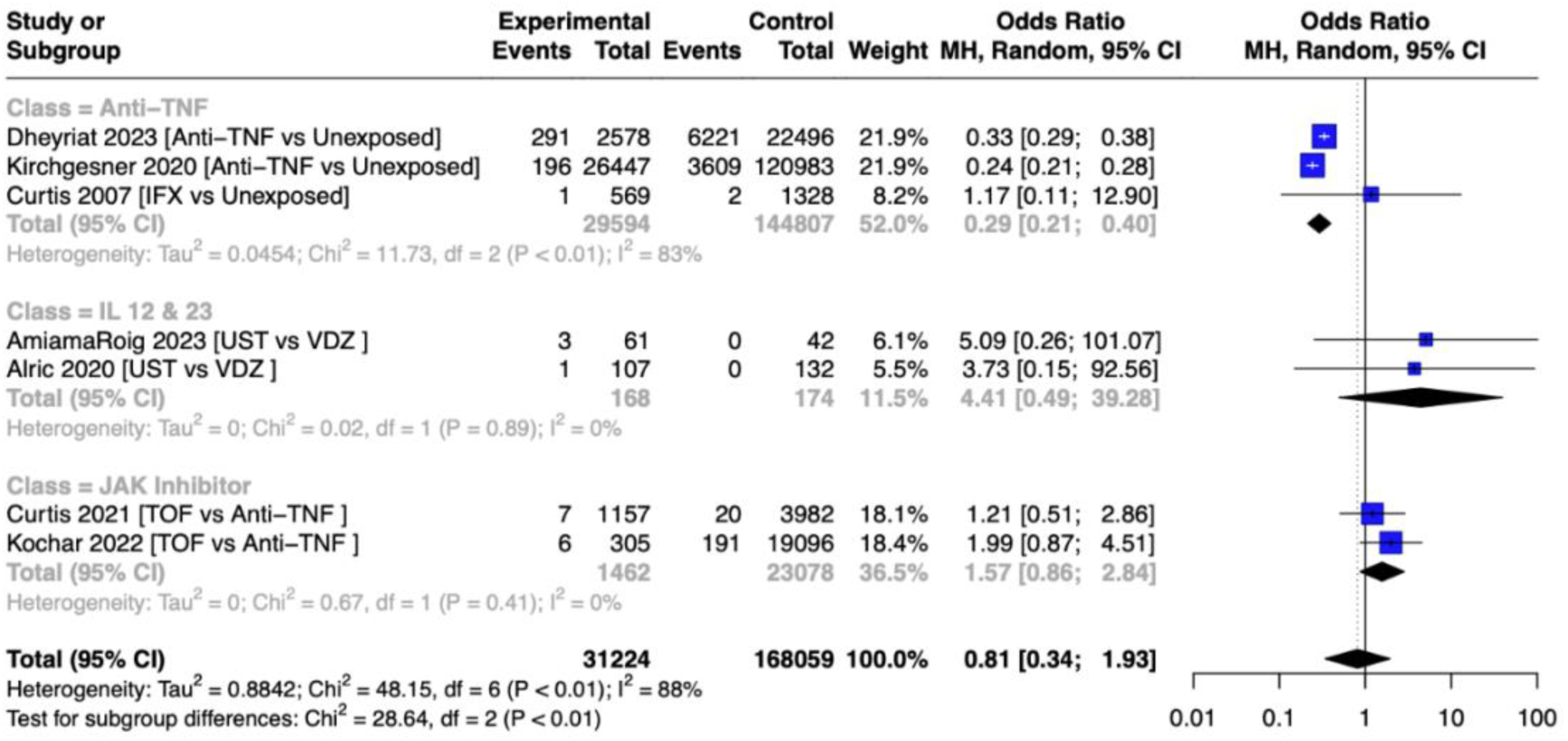
Forest Plot of Pooled Analysis for the MACE among Observational Studies.

### Risk of Bias Assessment

The risk of bias for RCTs was evaluated using the Cochrane RoB 2 tool, with 26 studies showing low risk and one study showing high risk. Observational studies were assessed using the ROBINS-I tool, with four studies judged to be at low or moderate risk, one at critical risk, and two at serious risk of bias in at least one domain. For detailed results of the assessment (see Supplementary Materials).

### Publication Bias

Visual inspection of funnel plot suggested a slightly asymmetry. However, Egger’s test (bias estimate -14.5, SE = 18.3, p = 0.23) showed lack of evidence on publication bias. The large standard error indicates considerable uncertainty in this assessment, possibly due to the limited number of studies (see Supplementary Materials, Table S2,3).

## Discussion

In this comprehensive meta-analysis, we synthesized data from forty-three studies, encompassing RCTs and observational studies, providing a broad perspective on the use of advanced therapies in IBD management and their potential cardiovascular implications. Overall, our findings indicate no significant difference in the risk of major adverse cardiovascular events (MACE) between patients receiving advanced therapies and those receiving placebo or conventional treatment.

In the primary analysis, we focused on direct comparisons between advanced therapies and placebo in RCTs and found a placebo-controlled RCTs showed a non-significant trend toward reduced MACE risk (OR, 0.60, 95% CI: (0.24-1.51)). The low heterogeneity (I² = 0%) suggests consistency across the included studies. However, the wide confidence interval highlights the uncertainty, likely due to the limited number of studies examining this specific outcome, short-term follow-up and the low event rate. To address the issue of failure to take the sample size of zero-event studies into consideration, we applied the Mantel-Haenszel (MH) method, which corrects for no events in one arm. However, this approach does not fully account for situations where both arms have zero events, potentially skewing the results. As a result, a sensitivity analysis was performed using the continuity correction method to adjust for zero-event studies. This method adds a small value (the reciprocal of the opposite treatment arm’s size) to zero cells, thereby balancing the comparisons and reducing bias in meta-analyses involving sparse data. The shift of the odds ratio toward the null after correction reflects the importance of properly accounting for zero-event studies when event rates are low.

When stratified by pharmacological class, IL-12/IL-23 inhibitors (OR 0.35; 95% CI: 0.05–2.21) and JAK inhibitors (OR 0.57; 95% CI: 0.16–2.06) showed non-significant trends toward reduced MACE risk, while anti-TNF therapies (OR 3.04; 95% CI: 0.31–29.47) demonstrated a non-significant trend toward increased MACE risk. Additionally, the secondary analysis, which included studies comparing higher versus lower doses, suggested potential dose-dependent effects for anti-TNF agents and JAK inhibitors, with notable differences between the induction and maintenance periods. Studies have demonstrated that higher therapeutic concentrations of advanced therapies are associated with improved clinical responses in IBD patients, especially during induction, further supporting the possibility of dose-dependent effects^62,63^. While the induction phase may carry a different cardiovascular risk than the maintenance phase, and variations in dosing and administration strategies could influence this risk, existing data do not allow for a definitive assessment. The relationship between disease activity and cardiovascular risk further complicates this evaluation. Patients with active IBD are at a greater risk for cardiovascular events, particularly during disease flares, while those in remission appear to have a risk comparable to the matched population^64^. A nationwide Danish study also documented a significantly increased cardiovascular risk immediately following an IBD diagnosis (incidence rate ratio [IRR]:4.6, 95% CI: 3.9–5.4), which gradually declined over time^65^. It remains unclear whether the elevated cardiovascular risk observed early in IBD reflects true disease-related risk, or other residual confounding.

The incorporation of long-term follow-up studies provided consistent results with the RCT findings, showing no statistically significant difference in MACE risk. For IL-12/IL-23 inhibitors, the pooled analysis revealed an OR of 0.25, 95% CI (0.06-1.10), suggesting a trend toward a reduction in MACE risk, indicating that the cardiovascular benefits of IL-12/IL-23 inhibitors may become more apparent over extended treatment periods. Whereas, for JAK inhibitors, the overall OR was 0.90, 95% CI (0.22-3.66), close to 1.0, suggests no clear risk reduction on MACE during long-term follow-up.

When it comes to observational studies, the apparent effects on MACE risk by pharmacological class differ from those observed in RCTs. It suggested a significant reduction in the risk of MACE for anti-TNF therapies, OR 0.29, 95% CI: (0.21-0.40), and a potential increased risk of MACE with JAK inhibitors, OR 1.57, 95% CI: (0.86-2.84), and IL-12/IL-23 inhibitors, OR 4.41, 95% CI: (0.49-39.28) have been demonstrated. Benefiting from the large sample size, several retrospective cohort studies demonstrated the statistically significant association. For instance, Dheyriat et al. (2023)^55^demonstrated a reduction in the odds of recurrent acute arterial events among IBD patients with a history of such events when treated with anti-TNF agents (OR 0.33, 95% CI: (0.29-0.38)). Similarly, Kirchgesner et al. (2020)^56^ reported a significant decrease in the odds of acute arterial events in IBD patients receiving anti-TNF therapy (OR 0.24, 95% CI: (0.21-0.28)).

It is important to note that RCTs typically exclude individuals with a pre-existing cardiovascular disease, whereas observational studies often include such patients. This key difference in study populations may partly explain inconsistent findings between two types of studies. Also, it is critical to acknowledge methodological limitations inherent to observational studies, particularly confounding by indication with a non-user design for the control arm given anti-TNF therapy is often prescribed to patients with refractory or severe IBD, who have a higher baseline risk of cardiovascular events than non-user. This systematic difference between treated and untreated groups complicates the interpretation of findings. Moreover, patients in the observational studies were older (mean age (SD), 66.2 (14.2) and 46.2 (16.3) for Dheyriat and Kirchgesner et al.) than those enrolled in the RCTs (37∼42 years old). To emulate randomization, propensity score and other types of matching or weighting approach have been applied, yet residual confounding remains a potential factor that might lead to these inconsistent results. For example, several major confounders, such as underlying CVD risk and the severity of IBD, were either not measured or were inappropriately accounted for in the matching process. Nevertheless, the high heterogeneity observed (I² = 88%), and small sample size (especially for studies where patients received IL-12/IL-23 inhibitors) indicates considerable variability across studies and emphasizes the need for careful interpretation. Finally, the current observational studies primarily included middle-aged and older adults with IBD, neglecting adolescents and young adults, an underrepresented population, thereby limiting the generalizability of the findings, as the age difference could contribute to these varying results, as older populations may have a higher baseline cardiovascular risk, potentially influencing the observed protective or harmful trends of the therapies.

Our meta-analysis aligns with recent findings by Shehab et al.(2023)^13^(OR 0.69, 95% CI: (0.26, 1.82)), showing no statistically significant difference in MACE risk between IBD patients treated with advanced therapies and placebo. However, our study employed a refined methodological approach, excluding studies that did not explicitly report or measure 5-point MACE outcomes, which could introduce measurement error and selection bias^66^. We analyzed data from 2007-2024, incorporating recent advances in IBD therapeutics, such as the IL-23 inhibitor Risankizumab. Our findings in the RCTs were different than the network meta-analyses by Mattay et al.(2024)^67^, where elevated risks of MACE have been associated with the anti-TNF therapies (OR 2.49, 95% CI: (1.14–5.62)), JAK inhibitors (OR 2.64, 95% CI: (1.26–5.99)) and anti-IL-12/23 (OR 3.15, 95% CI: (1.01–13.35)). The inconsistent results were jointly caused by different population where we focused on patients with IBD instead of broader scope of IMIDs, which may capture varying risk profiles, and more rigorous statistical approaches we employed (i.e., continuity correction).

Our meta-analysis possesses several notable strengths. The comprehensive approach, incorporating a wide range of study designs, provides a holistic view of cardiovascular risk, capturing both evidence from RCTs and real-world. Our stringent inclusion criteria, specifically the requirement for explicit reporting of 5-point MACE, enhance the reliability of our findings. Our specific focus on IBD, rather than the broader scope of IMIDs, allows for a more nuanced understanding of cardiovascular risk in this particular population. The secondary analysis that focused on studies with long-term follow-up, innovatively considered a relatively longer latent period for MACE and avoided the protopathic bias. The low heterogeneity (I²=0%) observed in our primary analysis suggests consistency across included studies, lending credibility to our findings. However, this meta-analysis has several limitations. A key challenge lies in the rarity of MACE events within the studied population, which contributes to low statistical power and wide confidence intervals, limiting the precision of our estimates. RCTs are not specifically designed or powered to investigate adverse events such as MACE, potentially resulting in an underestimation of the true association between these therapies and MACE. This limitation highlights the importance of integrating evidence from observational studies, which can provide a more comprehensive assessment of safety outcomes over extended periods and in broader, real-world populations. The varied design (i.e. re-randomization), follow-up of studies and potential differences in research protocols and implementation amplified the diversity of populations included in our analysis. While this heterogeneity enhances the generalizability of our findings, it also introduces challenges in their interpretation. Additionally, differences in the definition of MACE across studies complicate comparisons and contribute to variability in the outcome measurement. The potential for publication bias, though not statistically significant in our analysis, remains a concern due to funnel plot asymmetry and the non-trivial standard error in Egger’s test. This highlights the need for more inclusive studies, incorporating unpublished and negative results, to ensure robust conclusions for rare events like MACE.

It’s important to note that the relationship between advanced therapies and cardiovascular risk in IBD patients is complex and not fully understood. These therapies exhibit pleiotropic effects due to their complex interactions with the immune system, influencing multiple signaling pathways^68–70^. Consequently, their impact on cardiovascular risk factors and vascular function may vary based on individual patient characteristics, disease activity, and treatment duration. This complexity underscores the need for nuanced understanding in clinical application.

## Conclusion

In conclusion, we found no association between the use of advanced therapies and elevated MACE risk compared with using placebo or conventional treatments for IBD patients. While short-term studies showed no significant difference, the inclusion of long-term observational and real-world data highlighted a more favorable cardiovascular profile. The divergence between RCTs and observational studies underscores the challenges in evaluating MACE risk: RCTs are less biased but often limited by small sample sizes and short follow-up, whereas observational studies capture real-world effectiveness but are prone to bias, limiting causal inference. Future well-designed observational studies with long-term follow-up and comprehensive MACE endpoints are essential to clarify these associations and guide clinical practice.

## Acknowledgements

Amirah H. Alnahdi is supported by the Ministry of Health of Saudi Arabia and the Saudi Arabian Cultural Mission in the USA.

## Data Availability Statement

The data supporting the findings of this study are provided in the supplementary materials.

## Abbreviations

IBD: Inflammatory bowel diseases
CD: Crohn’s disease
UC: ulcerative colitis
IMIDs: immune-mediated inflammatory disorders
anti-TNF: anti-tumor necrosis factor
IL: interleukin
JAK: Janus kinase
S1PR: sphingosine-1-phosphate receptor
MACE: major adverse cardiovascular events
OR: odds ratio
RR: relative risk
HR: hazard ratio
RA: rheumatoid arthritis
axSpA: axial spondyloarthritis
CRP: C-reactive protein
TNF-α: tumor necrosis factor-alpha
RCT: randomized controlled trial(s)
LTF: long-term follow-up
RoB 2: Cochrane Risk-of-Bias Tool for Randomized Trials
ROBINS-I: Risk of Bias in Non-randomized Studies - of Interventions
CI: confidence interval
MH: Mantel-Haenszel method
I²: I-squared statistic
ADA: adalimumab
CZP: certolizumab pegol
ETZ: etrolizumab
FIL: filgotinib
IFX: infliximab
OZA: ozanimod
PBO: placebo
RZB: risankizumab
TOF: tofacitinib
UPA: upadacitinib
UST: ustekinumab
VDZ: vedolizumab
HIR: higher induction regimen
SIR: standard induction regimen
CA: clinically adjusted
TDM: therapeutic drug monitoring
TOT: treat-to-target
SOC: standard-of-care
ew: every week
eow: every other week
CVD: cardiovascular disease
IRR: incidence rate ratio

